# Quantifying the global burden of lead exposure from dietary lead intake

**DOI:** 10.64898/2026.07.20.26358457

**Authors:** Chris Kinally, Howard Hu, Richard Fuller

## Abstract

**Background:** Lead exposure is estimated to cause approximately 3.5 million premature deaths a year, yet the key ongoing sources of lead exposure are unclear.

**Methods:** We estimated the contribution of dietary lead intake to global blood lead levels (BLLs) for 7-year-old children and 22-year-old adults by applying the All-Ages Lead Model (AALM) to calculate blood lead levels (BLLs) based on 25 total diet studies (TDS) that quantify dietary lead intake across 46 countries.

**Results:** For children, the population-weighted average dietary lead intake in low- and middle-income countries (LMICs) (32.0 μg/day) was found to be more than three times higher than in high-income countries (HICs) (9.3 μg/day), and more than 10 times higher than the FDA’s reference level for children (2.2 μg/day). The average impact on BLLs for children is estimated to be near 29 μg/L in LMICs and near 12 μg/L in HICs. Averaged across the TDS data, vegetables (27%) and cereals (24%) were found to contribute the most to dietary lead.

**Conclusions:** While there are limitations associated with biokinetic modelling and the TDS data from LMICs, these results suggest that the contribution of dietary lead intake to global lead exposure is in the region of 40 to 50%, suggesting, in turn, that dietary lead intake is likely a major global driver of lead poisoning. Lead absorbed from the environment into food crops is expected to be the key driver of dietary lead. Current regulatory levels for maximum lead concentrations in foods (0.05-0.3 mg/kg) are out-of-date and may imply a dietary lead intake of 200 μg/day, far higher than the FDA reference level (2.2 μg/day). Collecting representative TDS data in high lead burden countries should be a priority. Further research is also recommended on upstream lead sources and pathways of lead uptake in plants, driving global food contamination.

This manuscript is a preprint and has not undergone peer review. It may be revised following review.

**Highlights:**

- Dietary lead intake is likely a major driver of global lead exposure
- Food contamination is largely driven by environmental pollution
- International reference levels for lead levels in food are out-of-date
- Levels in HICs show interventions to reduce dietary lead intake are possible
- Research is needed to aid interventions to reduce lead contamination in food crops

## 1. Introduction

Lead poisoning is estimated to cause between 3.5 to 5.5 million premature deaths and the loss of 765 million IQ points per year ^1 2^. The magnitude of the global lead health burden is well understood, and some of the most common sources of lead exposure are well documented ^3^, including contaminated consumer products, industrial pollution, environmental contamination, and occupational exposure. However, it remains unclear which sources and pathways of lead exposure contribute the most to lead health burdens at either a national or global scale, posing a challenge to the task of prioritizing mitigation strategies.

Dietary lead intake and absorption from contaminated food is known to be one of the key drivers of lead exposure; lead is absorbed from the environment into food crops and also transferred through the food chain into animal products ^4^. Even low-level lead contamination in staple foods risks substantial cumulative health burdens across populations due to chronic exposure. Both the World Health Organization (WHO) ^5^ and the European Food Safety Authority (EFSA) ^6^ have stated that food is the primary source of lead intake for non-occupationally exposed populations. However, the quantitative justification for these claims is unclear, especially in low- and middle-income countries (LMICs) where most of the global lead poisoning health burden is concentrated.

The first reference level for dietary lead intake was defined by the joint WHO and UN Food and Agriculture Organization (FAO) Joint Expert Committee on Food Additives (JECFA) in 1999, which defined a provisional tolerable weekly intake (PTWI) of dietary lead intake of 25 μg per kg bodyweight (μg/kg BW) of lead per week (i.e. 214 μg per day for a 60 kg adult) ^7^. However, JECFA since withdrew the PTWI in 2011 ^8^. More recently, in 2022, the US Food and Drug Administration (FDA) defined a substantially lower Interim Reference Levels (IRL) for dietary lead intake from the US of 2.2 μg/day for children and 8.8 μg/day for women of childbearing age, designed to reflect a BLL impact of 3.5 μg/L ^9^.

The FDA’s dietary lead intake IRLs were computed assuming a linear dose response relationship between chronic dietary lead intake and BLLs ^9^. The linear BLL conversion factors used by the FDA are 1.6 μg/L BLL increase per μg Pb ingested per day (μg/L per μg Pb/day) for children and 0.4 μg/L per μg Pb ingested/day for adults, based on empirical observations ^10^. JECFA proposed BLL conversion factors as ranges: 0.5–1.16 μg/L per μg ingested Pb/day for children and 0.23–0.7 μg/L per μg ingested Pb/day for adults ^11^. Hence, JECFA recognised uncertainties with these linear BLL conversion factors and assumed that the FDA’s conversion factor for children to be an upper bound estimate.

However, the relationship between BLLs and lead ingestion is not linear; BLL impacts have been shown to diminish at higher dietary lead intakes ^6^. Another approach to estimate the BLL impacts of dietary lead intake is with biokinetic modelling that employ computational models that consider how substances are absorbed, distributed, metabolised, and excreted throughout the body ^12^, considering age, sex, and other individual factors. Currently, the most advanced biokinetic model for lead is the All Ages Lead Model (AALM) ^12^, developed by the US Environmental Protection Agency (EPA). Several studies have validated the proposition that BLLs predicted by the AALM are in line with measured BLLs ^12^. Nonetheless, the accuracy of BLLs predicted with biokinetic modelling is dependent on the accuracy of the input data quantifying lead exposure – in this instance, the accuracy of dietary lead intake data.

Many governments have commissioned national Total Diet Studies (TDS), quantifying national average levels for the dietary intake of lead and other contaminants. TDS use a combination of diet surveys and food screening, aiming to determine population-representative average intake levels of different nutrients and contaminants. The largest TDS to date has been conducted by the European Union (EU), collecting over 280,000 food samples across 27 countries between 2013-2023 ^13^. Meanwhile, China produced their 6^th^ TDS in 2022, showing trends in national dietary lead intake since 1990 ^14^. Dietary lead ingestion rates are typically reported in μg of lead consumed per kg of body weight per day (μg/kgBW/day). In this sense, TDS data provide reliable estimates for national average levels of dietary lead intake at different ages.

In 2019, Carrington et al. ^15^ reviewed available TDS data to estimate the global burden of intellectual disability of dietary lead intake. Carrington et al. used the JECFA linear conversion factor to use TDS data to estimate blood lead levels (which were then used to estimate impacts on IQ), however TDS data were mostly restricted to HICs ^15^. Carrington et al. found a mean dietary lead intake of 21.1 μg/day for European children and estimated a BLL impact of 22.5 μg/L. Since 2015, the availability of dietary lead intake data has increased with the publication of many additional TDS and meta-analysis studies reviewing lead levels in food crops in LMICs and globally ^16–18^.

The aim of this study is to evaluate the contribution of food contamination to the global lead poisoning health burden using the most recent available TDS data to estimate average levels of dietary lead intake in both LMICs and HICs, and using biokinetic models to estimate associated BLL impacts for children and adults. Key foods and food groups driving national dietary lead intake are identified. Methods of quantifying BLL impacts from dietary lead intake are compared: AALM, IEUBK and the linear conversion factors from FDA and JECFA. Predicted BLL impacts from dietary lead intake are compared against estimates of national and regional average BLLs currently made by the Global Burden of Disease project of the Institute for Health Metrics and Evaluation ^9^. Finally, regulatory levels on maximum lead concentrations are discussed.

## 2. Method

### 2.1 Literature search

A semi-structured literature search was performed to collect TDS datasets that quantify dietary lead ingestion rates. TDS datasets were found using a combination of Scopus, Google Scholar and ChatGPT. Search strings included the keywords: “national”, “dietary”, “lead”, “intake”, “total”, “diet”, “study”. Snowballing was also used, yielding additional TDS datasets referenced within studies. Both peer-reviewed academic literature and grey literature (government-commissioned TDS reports) were considered. Continental, national and subnational datasets were included. In the cases where multiple national TDS studies have been conducted (EU, China, France, and Brazil), the most recent datasets were considered. Due to the limited availability of dietary lead intake data in LMICs, the inclusion criteria were deliberately inclusive to capture as much relevant data as possible. TDS dataset were included if they i) quantified estimates for dietary lead intake (μg/kgBW/day or μg/day) derived from pairing data for food consumption and lead concentrations in foods; ii) followed a recognisable TDS structure, commissioned by governments, citing the EFSA TDS methodology, or citing a peer-reviewed sub-national equivalent; (iii) provided sufficient detail on sampling to extract dietary lead intake rates for a defined population.

To account for the heterogeneity between the TDS datasets, national and sub-national studies are distinguished, studies reporting only adult intake data are distinguished (which systematically underestimates child intake, Section 3.1), and potentially confounding exposure pathways (tap water, adulterated spices, and artisanal cookware) are screened out where identifiable (Section 2.2).TDS datasets were considered to have a national scope if they were conducted by a government body, if the EFSA TDS methodology guidelines were cited, or if food samples were collected from multiple regions with a rationalised sampling structure. A total of 37 TDS datasets were found, of which 25 TDS datasets were included after accounting for countries where multiple national studies had been conducted. For each TDS data set, characteristics, including sampling scope, date, age groups covered, dietary lead ingestion rate data, analytical techniques used, how samples below the limit of detection were handled, and any adjustments applied are provided in the Supporting Information. Finally, the limited available dietary lead intake data in LMICs were compared against supporting evidence from Xiao et al.’s global meta-analysis on lead levels in cereal crops ^16^, which reports dietary lead intake levels from cereal crops – a key driver of dietary lead intake in LMICs. In analysing the data, distinctions were made between HICs and LMICs (based on current World Bank criteria) and between national and sub-national studies. Upper-middle-income countries were included as LMICs (Brazil and Argentina). The limitations of this inclusive approach and the lack of nationally representative TDS data in LMICs are discussed in Section 4.1.

### 2.2 Dietary lead intake data

Dietary lead ingestion rates were extracted from the TDS datasets, commonly reported as μg Pb per kg body weight per day (μg/kgBW/day). For studies that report dietary lead intakes as lower and upper bound estimates, the average between the upper and lower bound estimates was used. Dietary lead ingestion rates were converted to μg/day for each year of age for the purpose of modelling BLL impacts. Average body weights are defined, taken from the WHO Weight-for-age charts for 0-10 years old (3.3-31.5 kg), averaged for both sexes and then assuming a linear increase in body weight from 10 years old (31.5 kg) to 20 years old (60 kg). Body weight is assumed to remain constant from 20 years old at 60 kg (the default adult body weight used by the FAO and WHO ^19^). Some (9/25) of the TDS datasets quantified different dietary lead intake rates (μg/kgBW/day) for different age groups, which is included in our analysis. These studies all show dietary lead intake rates per unit body weight (μg/kgBW/day) for children to be higher than adults. For the datasets that did not provide dietary lead intake data for children, the adult dietary lead ingestion rates were assumed for all ages, which is therefore expected to underestimate dietary lead intake for children. For each TDS dataset, the dietary lead intake data (μg/kgBW/day and μg/day) for 1-25 years old used for biokinetic modelling are provided in the Supporting Information. Dietary lead intake for children under 1 (who may not be eating solid food) is not included in our analysis (assumed to be zero) because of the lack of data on lead levels in infant formula and breastmilk. Where available, dietary lead intake data per food group were extracted from each TDS dataset.

The TDS datasets were screened to mitigate potential contributions to dietary lead intake from tap water, adulterated spices and artisanal metal cookware. Tap water and drinking water were removed from 8 of the TDS datasets (shown in the Supporting Information). For other datasets, water was not included or did not have detectable levels of lead. Contributions from spices were removed from one dataset ^20^ because high lead levels were flagged, suspected from dust contamination whilst airdrying spices. None of the other TDS studies found lead concentrations in spices > 1 mg/kg. Contributions from cookware were considered negligible in HICs. For the 14 LMIC TDS, eight of the studies (indicated in the Supporting Information) did not provide enough detail on the method of food preparation to rule out potential contributions from metallic cookware. However, as TDS studies are expected to be carried out in laboratories, the risk of contamination from artisan metal cookware is assumed to be low.

### 2.3 Blood lead level impacts

The BLL impacts associated with the TDS dietary lead intake rates are quantified using the AALM version 3.0 64-bit (released March 2024). For the AALM inputs, 0-25 years old were considered, with food the only active media, linearly interpolated exposure was selected, and non-linear red blood cell (RBC) calculation was selected (the default), allowing for RBC saturation, and the default value for bioavailability was used. For each TDS, the dietary lead intake μg/day data were entered for each year of age (0-25 years old), as shown in the Supporting Information. The BLL simulation was run for both males and females, and the BLL outputs were averaged.

The results of the AALM models were compared with those generated by the Integrated Exposure, Uptake, and Biokinetic (IEUBK) model (version 2.0; released May 2021); and the linear dietary lead intake-to-BLL conversion factors from the FDA and JECFA. Diet was the only lead ingestion pathway used; all other lead inputs (air, water, soil/dust, and maternal) were set to zero. The IEUBK model provides BLL predictions as an average value for both sexes. For the FDA, a 1.6 μg/L BLL increase per μg of lead consumed per day was considered for children, and a 0.4 μg/L BLL increase per μg of lead consumed per day for adults ^9^. For JECFA, the midpoint, upper and lower bounds were considered: 1.05, 0.5 and 1.6μg/L per μg Pb/day, for children, and 0.465, 0.23 and 0.7 μg/L per μg lead consumed per day for adults, respectively ^11^.

To estimate the % contribution of dietary lead intake to the total burden of lead poisoning in LMICs and HICs, we used the cumulative population BLL lead burden metric ^21^: the average BLL impact from dietary lead intake in LMICs and HICs is divided by the average BLLs in LMICs and HICs, respectively. The lead burden for 7-year-old children and 22-year-old adults was estimated separately for each country. To estimate the average BLL impacts from dietary lead intake across LMICs and HICs, a population-weighted arithmetic mean was used, considering the national average BLL impact predicted from each TDS study and the national population of each age group. National population and regional average BLL data (by age group) were obtained from the Institute for Health Metrics and Evaluation data repository ^22^. For children, the IHME 5-9 years old age group category was considered. For adults, the IHME 20-24 years old category was considered.

## 3. Results and Discussion

### 3.1 Scope of TDS data and headline results

The 25 selected TDS were published between 2005 to 2025. The sampling scope, dietary intake values, and predicted BLL impacts are shown for each TDS dataset in the Supporting Information. Thirteen of the TDS were considered as being national in scope (section 2.1); the remaining 14 were sub-national. Of the TDS, 15 were from LMICs, and of these, 4 had a national scope. Most TDS quantified dietary intake for adults, whereas only 9 studies gave specific intakes (per kg BW) for children. Regarding the TDS that quantified dietary lead intake for both children and adults, children were uniformly found to have significantly higher rates of dietary lead intake per unit bodyweight (μg/kgBW/day). Notably, all of the national-scale studies that quantified specific dietary lead intake rates for children were in HICs. The remaining 17 studies only give dietary lead intake rates for adults, values that could be expected to underestimate the dietary lead intake for children based on the results of the TDS mentioned above that quantified dietary lead intake for both children and adults. Two of the studies (US and Argentina; ^23,24^) only reported dietary lead intake for children, not adults. These two studies were not used in our analysis to estimate dietary lead intake rates for adults because they may overestimate adult lead intake. The dietary lead intake and associated BLL impact for each individual TDS is provided in the Supporting Information; the population-weighted average across all TDS in HICs and LMICs is shown for children and adults in Table 1.

**Table 1.**
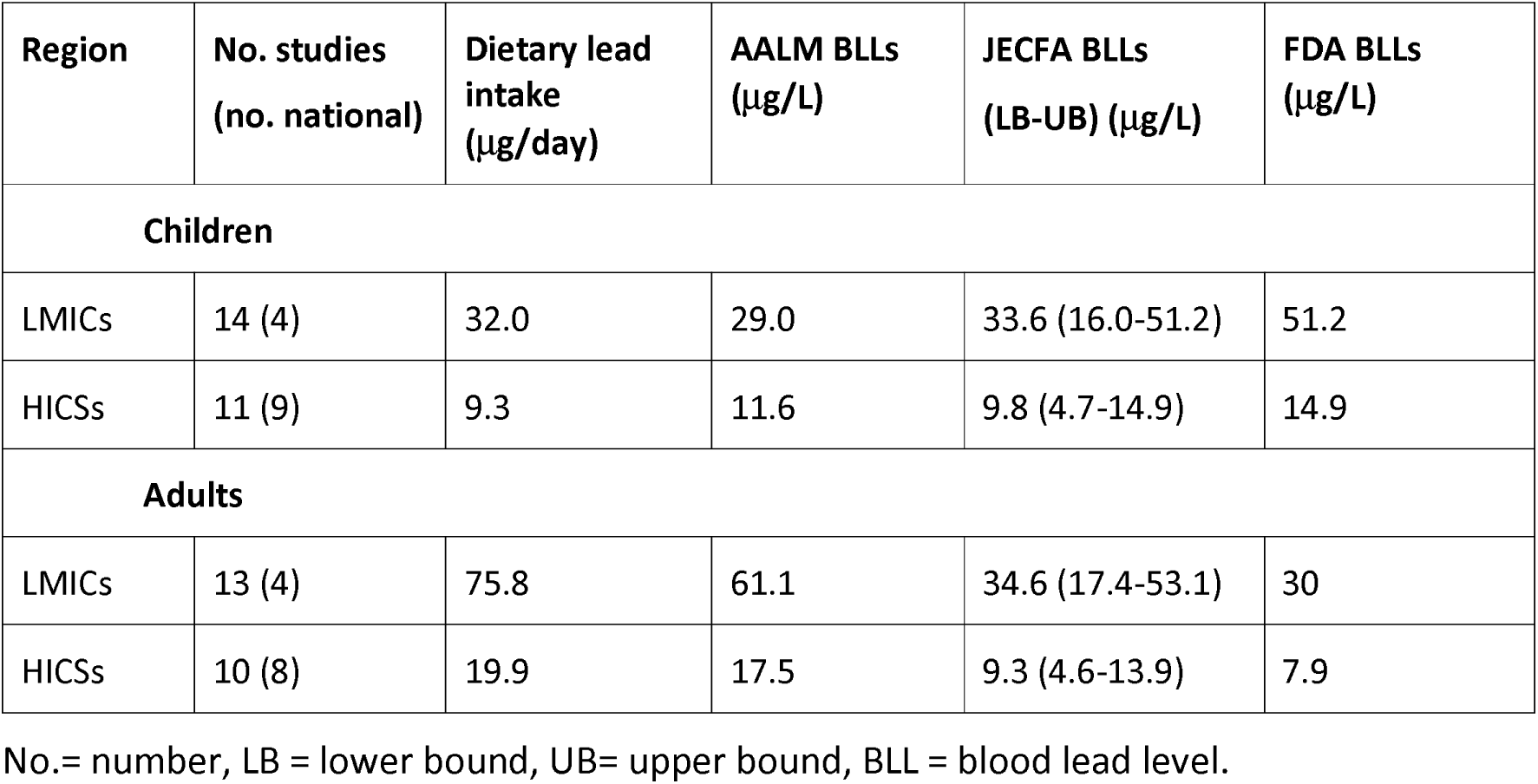
Population-weighted mean dietary lead intake for 7 year old children (22.6 kg) and 22 year old adults (60 kg) across all TDS studies and BLL impacts estimated by the AALM, JECFA, and FDA conversion factors.

### 3.2 Dietary lead intake

Dietary lead intake (μg/day) for each TDS is shown for 22-year-olds in Figure 1, and for 7-year-olds in figure S1 in the supporting information. The population-weighted mean dietary lead intake was substantially higher in LMICs (32 μg/day for 7-year-old children) than in HICs (9.3 μg/day for 7-year-old children). However, even in HICs, the average dietary lead intake for children (9.3 μg/day) is still four times the FDA reference level for children (2.2 μg/day). Across all TDS studies, none were below the FDA’s IRL for children, and only two of the TDS (Ireland and UK) were below the IRL for women of childbearing age (8.8 μg/day). There is significant variation in dietary lead intake between countries, and no clear trend in dietary intake by year of publication. However, trends in reducing dietary intakes can be observed within countries where multiple TDS studies have been conducted ^25^.

**Figure 1.**
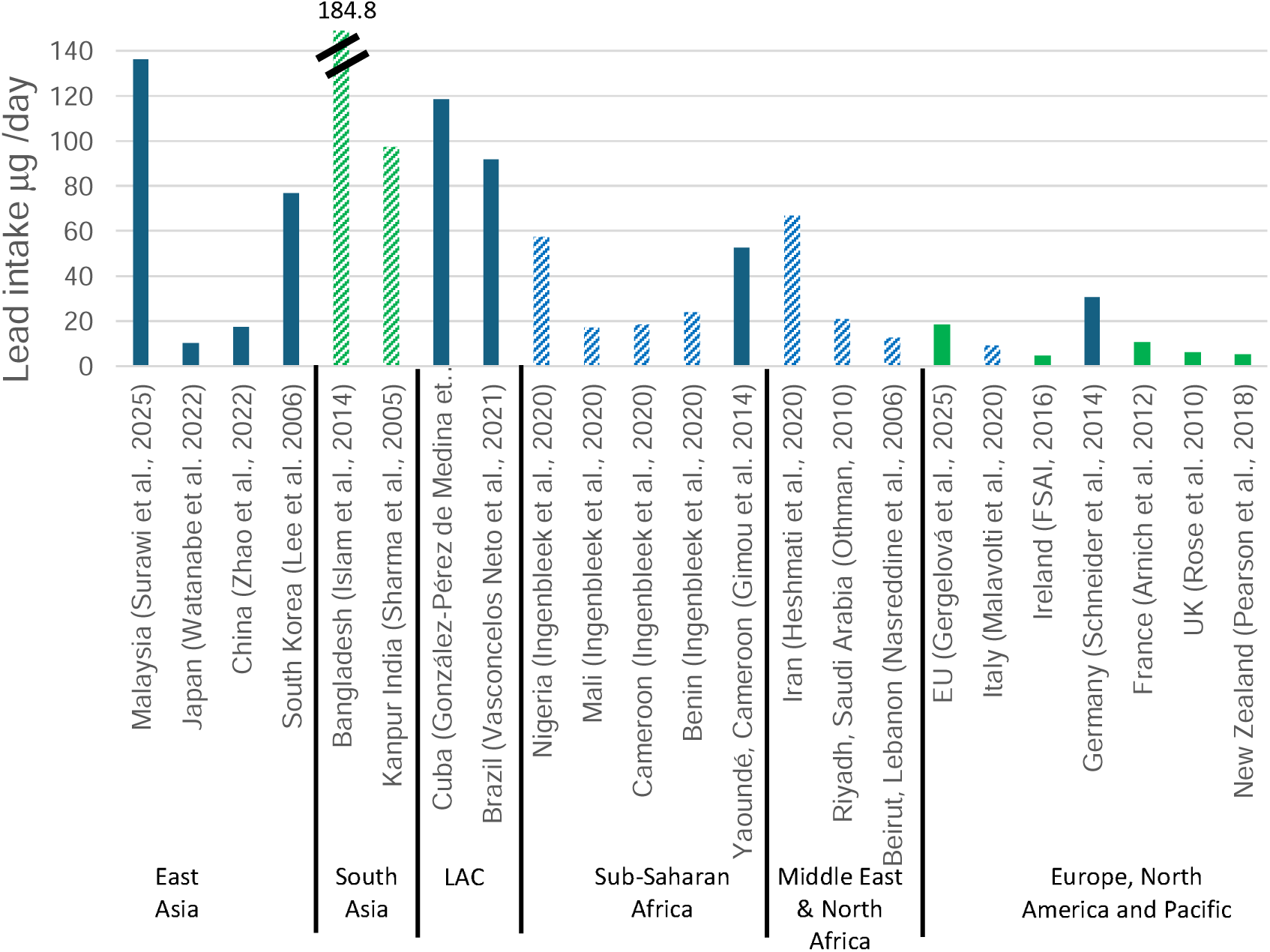
Dietary lead intake (μg /day) for 22-year-old 60 kg adults. Solid fill = national data, pattern fill = sub-national datasets, green = child specific dietary lead intake data. LAC = Latin America & Caribbean

For adults, five of the TDS exceed 80 μg/day: Malaysia ^26^, Bangladesh ^27^, India ^28^, Cuba ^29^ and Brazil ^30^. Three TDS exceed 100 μg/day for adults (Malaysia, Bangladesh and Cuba), with the highest recorded lead intake being 184.4 μg/day in Bangladesh. The high dietary lead intake in the subnational TDS in Bangladesh (184.4 μg/day for adults) ^27^ appears to be an outlier. However, Bangladesh (and South Asia, generally) is known to be a hotspot for food crop contamination, and the TDS data is in line with Xiao et al.’s recent meta-analysis, which estimates a dietary lead intake >150 μg/day for adults from cereal crops alone ^16^ (discussed in Section 4.1). Across all of the TDS data for Europe, North America and the Pacific, dietary lead intake is less than 20 μg/day for adults, except for Germany (31 μg/day).

Comparing the available TDS data for children is skewed by the key distinction between the TDS studies that collected specific lead intake data for children (shown in green in Figure 1) and the studies which only collected lead intake data for adults (shown in blue in Figure 1). All of the studies that quantified rates of dietary lead intake for children found children to have higher rates of lead intake per unit body weight. For example, for 7-year-olds, the rate of lead intake (0.8 μg/kgBW/ day in the EU, 7 μg/kgBW/day in Bangladesh) can be more than double the rate of lead intake for adults (0.36 μg/kgBW/day in the EU, 3.08 μg/kgBW/day in Bangladesh). Therefore, our results are expected to significantly underestimate dietary lead intake for children for the majority of the TDS studies which only report dietary lead intake data for adults, potentially understating by as much as 50% of the actual value in some cases.

### 3.3 Lead Intake by food group

Sixteen of the 25 TDS quantified dietary lead intake by food group. Relative (%) dietary lead intake per food group is shown in Figure 2, and total intake per food group (μg/day) is shown in Figure S2 in the supporting information. Averaged across all the studies, vegetables (27%) and cereals (24%) were the largest contributors to dietary lead intake, suggesting that lead absorbed into food crops from the environment is the primary pathway for dietary lead intake, accounting for around half of the total (Table 2). Of the other contributors to dietary lead intake, beverages (10%), milk and dairy (6%), meat (6%), and fruits (5%) account for the bulk of the remainder. When evaluating dietary lead intake from foods, both the lead concentration of the food and the quantity of food eaten must be considered. Generally, vegetables (particularly leafy greens) have a higher affinity to absorb lead but are eaten in lower quantities, whilst cereals have a lower affinity to absorb lead but are eaten in higher quantities. Lead contamination in other food categories, such as animal products and beverages, may also be explained by environmental uptake, as environmental lead pollution embeds into ecosystems and can accumulate in food chains.

**Figure 2.**
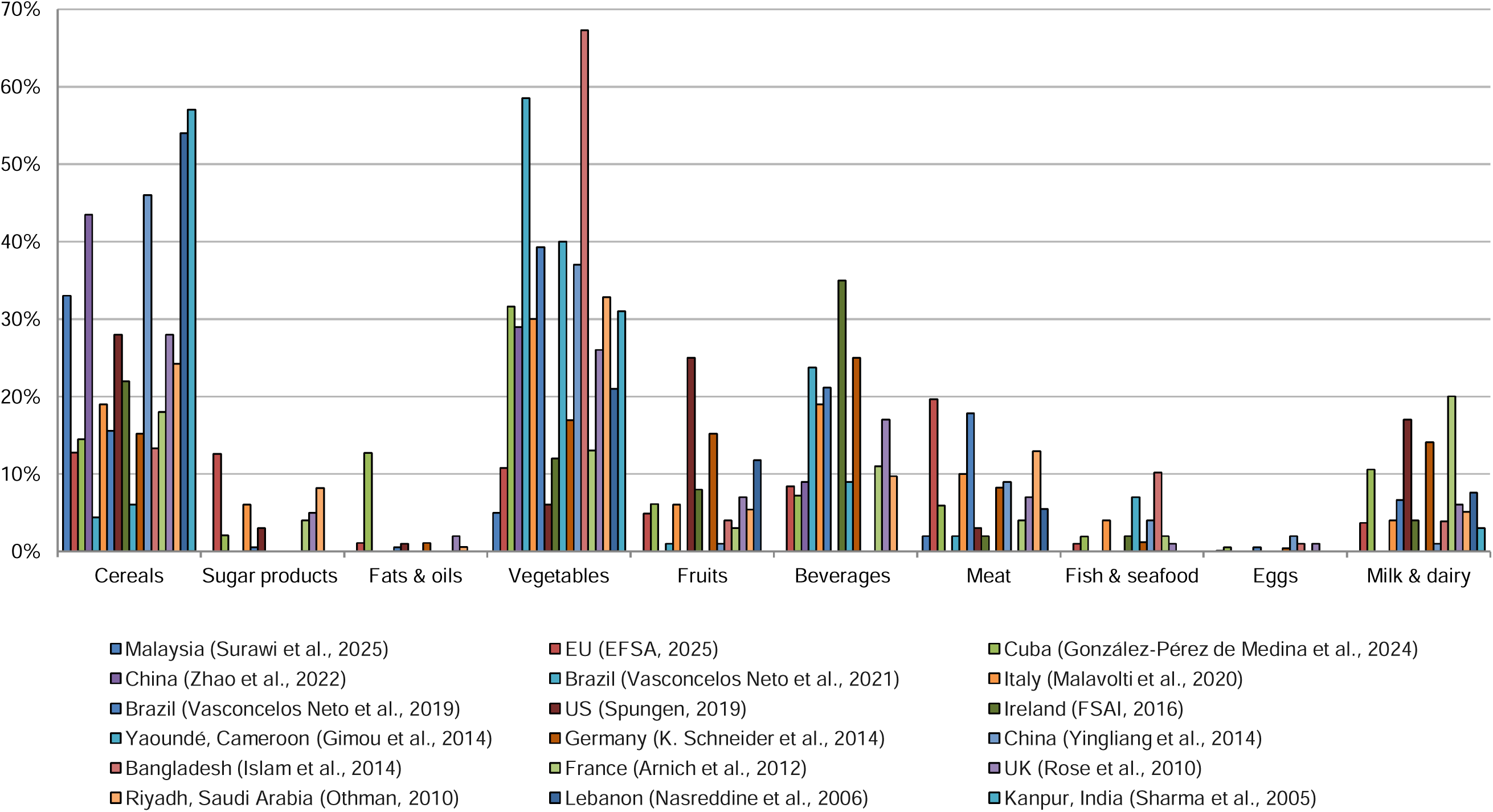
Relative dietary lead intake by food group (%) for the 18 TDS studies which provide data by food group. Where data are available, nuts are shown to contribute less than 1% to total dietary lead intake. Additional meta-analysis studies included for China 2014 and Brazil 2019, not included in the BLL impacts assessment (superseded by government commissioned national TDS). All beverages = alcoholic and non-alcoholic beverages, excluding tap wate

**Table 2.** Contribution of food groups to dietary lead intake, unweighted arithmetic mean and recorded range of % per food category across the 18 TDS studies which provide data by food group (see Figure 2).

| Food group | Average contribution to dietary lead intake (range) (%) |
| --- | --- |
| Vegetables | 27% (5-67%) |
| Cereals | 24% (4-57%) |
| Beverages (alcoholic and non-alcoholic) | 10% (0-35%) |
| Milk & dairy | 6% (0-20%) |
| Meat | 6% (2-20%) |
| Fruits | 5% (1-25%) |
| Sugar products | 2% (1-13%) |
| Fish & seafood | 2% (0-10%) |
| Fats & oils | 1% (0-13%) |
| Eggs | <1% (0-2%) |

Beverages (excluding tap water, see Section 2.2) averaged 10% of dietary lead intake. In Brazil, where lead intake from beverages was the highest (21.8 μg/day) ^30^, the beverages included in the TDS were fruit juices, coffee, tea and herbal teas. Across the 5 studies that specified lead intake from alcoholic beverages, alcohol showed a significant contribution to relative dietary intake (4-28%), although only in countries where total dietary lead intake is very low. The highest recorded dietary intake from alcohol was 1.5 μg/day in Italy ^33^, attributed to wine. All animal products combined contributed to only 15% of dietary lead intake – lead transfer through the animal food chain is inefficient, partly because lead accumulates in bones more than in edible tissues ^34^. However, eating wild game shot with leaded ammunition is shown to substantially increase dietary lead intake ^13^. The categorisation of nuts differed between some of the studies, either included in the fruit food group or vegetable food group. However, in the TDS where specific data for nuts is available, nuts are shown to contribute less than 1% to the total dietary lead intake (shown in the supporting information), so this discrepancy is not expected to impact the narrative of the results.

There are substantial variations between countries for the dietary lead intakes (μg/day) from individual food groups. Vegetables and cereals are the only categories to exceed 25 μg/day, recorded to exceed 25 μg/day in eight countries. The highest intakes were 124 μg/day from vegetables in Bangladesh ^27^, 53.7 μg/day from vegetables in Brazil ^35^ and 68.3 and 55.6 μg/day from cereals in Malaysia ^26^ and India ^28^, respectively.

The variation in dietary lead intake by food group may be explained by the combination of i) variation between food crops in their affinity to absorb lead from the environment, ii) local factors influencing lead uptake in food crops (such as sources of lead pollution and soil chemistry, etc.), and iii) variation between countries in the quantities of different foods eaten. Significant variation could also be caused by differences in how composite foods are defined between TDS, for example, if pastries are categorised as a sugar product or as cereals.

Lead contamination from post-harvest processing and cooking practices (such as artisanal aluminium cookware) also potentially contributes significantly to dietary lead intake. However, the TDS in Bangladesh ^27^, Malaysia ^26^ and India ^28^, which show extremely high dietary lead intakes (97.5-184.8 μg/day for adults), considered only raw foods (Bangladesh and India) or specified the use of stainless steel cookware (Malaysia), mitigating potential contributions from cookwareBLL impacts.

#### 3.3.1 Comparing methods of predicting BLL impacts

Methods of predicting BLL impacts from dietary lead intake are compared in Figure S3 in the supporting information. For children, the BLL impacts predicted by the AALM are well aligned with the IEUBK and the linear conversion factors from the FDA and JECFA. For 7-year-olds, the AALM is more conservative, showing lower BLL impacts than IEUBK and the FDA. At 6 years old (age limit for FDA and JECFA), the AALM (22 μg/L) is approximately at the midpoint between JECFA’s upper and lower bounds (25 μg/L). Whereas for 22-year-old adults, the BLL impacts predicted by the AALM (51 μg/L) are significantly higher than both the FDA (25 μg/L) and the midpoint between JECFA’s upper and lower bounds (29 μg/L). Notably, the FDA and JECFA linear conversion factors do not account for BLL impacts from cumulative dietary intake – BLL estimates only consider current daily lead intake. This is illustrated in Figure S3; after 20 years old, when dietary lead intake is assumed to remain constant at 75.8 μg/day, there is no change in BLL despite cumulative exposure. Whereas the AALM is more sophisticated and considers the impact of cumulative lifetime lead exposure.

We have higher confidence in the AALM results for children, as the results are relatively conservative compared to the consensus between the IEUBK model and the FDA and JECFA conversion factors. For adults, it is possible that the AALM potentially overestimates BLL impacts; further research is needed to increase the transparency of the BLL impacts of cumulative dietary lead intake.

#### 3.3.2 National BLL impacts

The precited BLL impacts for each TDS are shown for adults in Figure 3 (FDA conversion factor results), and for children in Figure S4 in the supporting information, compared against IHME’s estimates for national average BLLs. The FDA conversion factor is used to show the BLL impacts from diet for adults because it is more conservative than the AALM for adults (section 3.4.1). In-line with dietary lead intake, the predicted average BLL impacts in LMICs (32 μg/L for 7-year-olds) are at least double the precited average BLL impacts in HICs (9.3 μg/L for 7-year-olds, Tables 1 and 2). Whilst the BLL impacts are lower in HICs, dietary lead intake is expected to be the primary driver of BLL in both HICs and LMICs (section 3.5).

**Figure 3.**
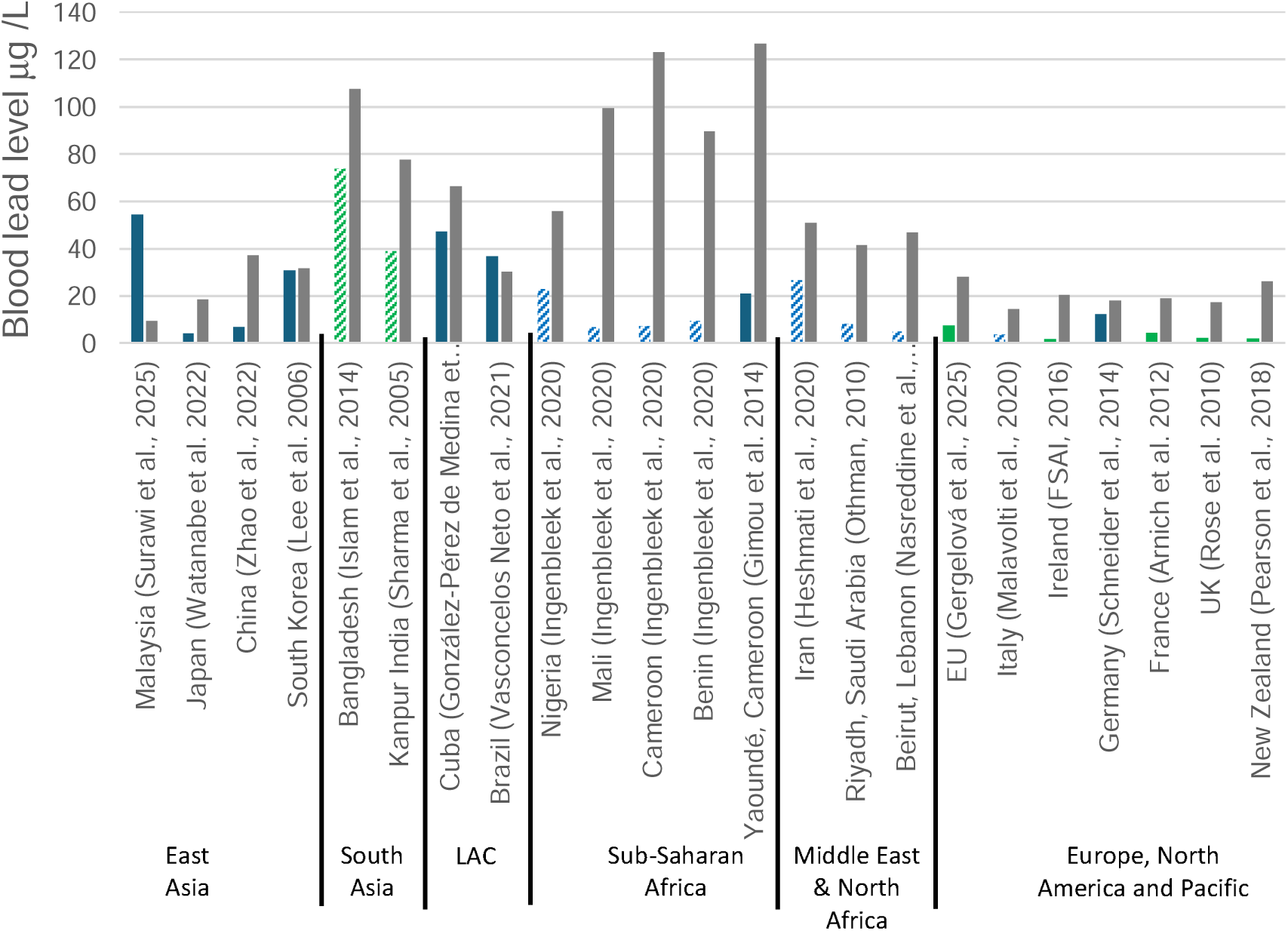
Blood lead level impacts (μg /dl) for a 22-year-old 60 kg calculated using the FDA’s linear conversion factor (0.4 μg/L per μg/day). Pattern fill = sub-national datasets, green = child specific dietary lead intake data, grey = IHME national average BLL for year of TDS publication, for TDS published after 2023 IHME 2023 (latest available) data is used. LAC = Latin America & Caribbean.

In a few cases, the predicted BLL impacts from dietary lead intake overshoot IHME’s estimated national average BLL. This overshoot suggests that either i) the TDS data overestimate dietary lead intake, ii) the AALM overpredicts BLL impacts, or iii) IHME underpredict national BLLs (discussed in limitations, section 4). There is significant uncertainty between different methods for predicting BLL impacts, although the AALM and FDA conversion factors are believed to be more conservative methods for children and adults, respectively, as shown in section 3.4.1. Moreover, there is significant uncertainty in IHME’s BLL estimates due to the lack of nationally representative BLL screening studies in LMICs. IHME rely on covariates relating to socio-demographics and time since leaded gasoline phase-out to predict national BLLs, not accounting for available TDS data on dietary lead intake. Hence, IHME’s national BLL estimates are expected to be less accurate for countries where BLL data is less available.

Malaysia, Brazil and Bangladesh are notable examples of the BLL impacts from dietary lead intake exceeding the IHME’s estimated national average BLL. The largest overshoot is shown for Malaysia, where the predicted BLL impact (49 μg/L for 7-year-olds) is more than 6 times IHME’s estimated national average BLL (7.7 μg/L for 5–9-year-olds in 2023). Child BLL data in Malaysia is limited, but the most recent study (2017) shows average BLLs between 69-85 μg/L (n=111) ^38^, and earlier data shows a mean of 40 μg/L for primary school children (n=346) in 2000. Hence, it is plausible that IHME substantially underestimate national BLLs for Malaysia. In Brazil, the national average dietary lead intake (1.53 μg/kgbw/day) ^30^ is similar to the level found by a separate meta-analysis on lead levels in food in Brazil (1.59-1.83 μg/kgbw/day) ^17^. Finally, whilst the sub-national TDS in Bangladesh appears to be an outlier with a very high dietary lead intake of 185 μg/day for 22-year-olds, Xiao et al.’s recent meta-analysis suggests adults in Bangladesh have a dietary lead intake >150 μg/day from cereal crops alone ^16^ (discussed in Section 4.1).

### 3.4 Share of global lead burden attributable to diet

The population-weighted mean BLL impacts in HICs and LMICs are compared against IHME’s 2023 estimated average BLL for 7-year-old children and 22-year-old adults in Table 3, respectively. For children, where we have higher confidence in the AALM results (see section 3.4.1), the BLL impact from diet predicted by the AALM are 48% of the expected average BLL in LMICS, with JECFA’s lower and upper bounds suggesting a range of 27-85%.

**Table 3.** Population-weighted arithmetic mean BLL impacts for 7 year old children (22.6 kg) and 22 year old adults (60 kg) across all TDS studies compared to estimates for regional average BLLs.

| Region | No. studies (national) | AALM BLL µg/L | FDA BLL µg/L | JECFA BLL (LB-UB) µg/L | IHME 2023 average BLL (20-24 y/o) µg/L | AALM BLL / IHME (%) | FDA BLL/ IHME (%) | JECFA BLL (LB-UB) / IHME (%) |
| --- | --- | --- | --- | --- | --- | --- | --- | --- |
| **Children** |  |  |  |  |  |  |  |  |
| LMICs | 14 (4) | 29.0 | 51.2 | 33.6 (16.0-51.2) | 60.0 | 48% | 85% | 56% (27-85%) |
| HICs | 11 (9) | 11.6 | 14.9 | 9.8 (4.7-14.9) | 14.1 | 82% | 106% | 70% (33-106%) |
| **Adults** |  |  |  |  |  |  |  |  |
| LMICs | 13 (4) | 61.1 | 30.3 | 34.6 (17.4-53.1) | 70 | 87% | 43% | 50% (24-76%) |
| HICs | 10 (8) | 17.5 | 7.9 | 9.3 (4.6-13.9) | 16.6 | 105% | 48% | 56% (28-84%) |

For adults, where the AALM may overestimate BLL impacts from diet, the AALM suggest 87% of BLLs in LMICs. Meanwhile, the FDA and JECFA conversion factors are more conservative, suggesting 43% and 50%, respectively (24-76% LB-UB). Hence, whilst the accuracy is limited by significant uncertainties associated with TDS data, biokinetic modelling and IHME’s estimates (discussed in section 4), the available data suggests that dietary lead intake is likely a major, and potentially the single largest, driver of BLLs in LMICs, potentially attributable to 40-50% of BLLs.

In HICs, the % contribution of diet to BLLs is expected to be higher than in LMICs for both adults and children. The AALM suggests 82% for children, and the FDA and JECFA conversion factors suggest 48% and 56%, respectively (28-84% LB-UB). Hence, dietary lead is also expected to be a major driver of BLLs in HICs, where TDS data is expected to be more representative.

### 3.5 Reference levels for lead in foods

Despite the results suggesting significant BLL impacts from dietary lead intake, the recorded lead concentrations in foods were below global regulatory standards in many studies. International maximum levels (MLs) for lead in food are defined by Codex standards ^39^, commonly 0.05–0.3 mg/kg (fresh weight) for many fruit and vegetable categories, 0.4 mg/kg for rice and 0.2 mg/kg for wheat. Similar category-based MLs have been adopted and are legally enforceable in many countries, such as the EU ^40^, Canada ^41^, China ^42^ and India ^43^.

These defined MLs are insufficient to prevent high BLL impacts from dietary lead intake. For example, in India, Sharma et al. ^28^ recorded adults to have a lead intake of 60 μg/day from cereal crops, consuming 500 g of wheat and rice per day at a mean lead concentration of 0.12 mg/kg. Despite the lead level in the cereal crops being below the Codex ML, the 60 μg/day lead intake is expected to cause a BLL impact of 24 μg/L for adults, from cereal crops alone. It’s feasible that a child’s dietary lead intake could exceed the FDA reference level of 2.2 μg/day by more than 100 times whilst consuming food that adheres to the Codex MLs; by eating 1kg of food at 0.22 mg/kg.

Similar to the FDA’s approach, MLs for lead in food should consider total daily dietary lead intake ^9^.

## 4. Limitations & areas for future research

The results suggest that dietary lead intake is potentially the primary global driver of lead poisoning, in line with previous statements from the WHO and EFSA (EFSA, 2010; WHO, 2007). However, there are significant uncertainties in the modelling of this study, specifically with i) the representativeness of TDS data, ii) the accuracy of predicting BLLs with biokinetic modelling, and iii) estimating the burden of lead exposure by comparing predicted BLLs with IHME estimates.

### 4.1 Lack of reliable TDS data in LMICs and limited geographic resolution

A key limitation for studies estimating the global health burdens of sources of lead exposure is the lack of representative data in LMICs, where the majority of the global lead burden is contained ^37^. Hence, similar to prior studies, this study relies on extrapolating the best available yet limited data (14 TDS studies in LMICs) across all LMICs.

Considering the lead burden index (cumulative population blood lead levels: average BLL multiplied by population size ^21^), 48% of the global lead burden is contained within the top five countries, which are all in Asia: India (19.5%), China (13.6%), Pakistan (8%), Bangladesh (3.8%), and Indonesia (3.2%) (considering IHME 2023 all ages data) ^21^. Meanwhile, Xiao et al. also show Asia (particularly South Asia) to be the current global epicentre for cereal crop contamination (a key driver of dietary lead intake, section 3.3), driven by industrial lead emissions. Xiao et al. specifically highlight India (median 1.02 mg/kg in corn), Pakistan (median 0.4 mg/kg in rice) and Bangladesh (median 0.31 mg/kg in corn) for having very high levels of food crop contamination, expecting adults’ dietary lead intake to exceed 120 μg/day in all three countries, from cereal crops alone. In agreement with Xiao et al., our results show the highest lead burdens (BLL impact from food multiplied by population size) in South Asia (India and Bangladesh), although relying on data from sub-national (TDS) studies.

Therefore, we expect the burden of lead exposure from dietary lead intake to be more concentrated in Asia, the global epicentre for both lead poisoning and food contamination. However, the lack of nationally representative TDS data conceals the extent of lead exposure from dietary intake in this region. Meanwhile, extrapolating available data from Asia may also potentially overestimate dietary lead intake in other regions, such as sub-Saharan Africa. In general, the health burden of dietary lead intake is expected to be substantial, so conducting nationally representative TDS in LMICs to improve the accuracy and reliability of these projections should be a priority.

Another key source of uncertainty is the rate of dietary lead intake for children. Only nine of the 26 TDS report lead intake rates for children, all of which found the lead intake rate for children (relative body weight, μg/kgBW/day) to be significantly higher than for adults. For example, Europe’s TDS found the rate of dietary lead intake for children (0.76 μg/kgBW/day, 3-9 years olds) to be more than double the rate for adults (0.31 μg/kgBW/day, >18 years old) ^13^. Hence, by relying on data for lead intake rates for adults, this study is expected to significantly underestimate dietary lead intake for children for the remaining majority (17/26) of the TDS. For future TDS investigating dietary lead intake in LMICs, collecting specific data for children should be a priority.

### 4.2 Biokinetic modelling

The accuracy of predicting the BLL impacts of dietary intake with biokinetic modelling is also limited. The AALM is built on available biokinetic data for HICs and may be less accurate in LMICs ^44^. A key area of uncertainty is the bioavailability of lead in food, which can vary between foods and based on dietary and nutritional context, with lead uptake suggested to increase with nutritional deficiency ^45^. We considered the default setting for bioavailability in the AALM, although we encourage further research to scrutinise the validity of the AALM’s assumptions for bioavailability in LMICs. Recognising the uncertainties of predicting BLL impacts from dietary lead intake, we review different methods for predicting BLL impacts: the AALM, IEUBK model, and conversion factors from JECFA and the FDA. The different methods show a broad range of predicted BLL impacts, but all suggest that dietary lead intake could be responsible for more than 40% of BLLs in LMICs.

### 4.3 Lead burden attribution with IHME estimates

Estimating the fraction of BLLs attributable to dietary lead intake in LMICs and HICs requires comparing the expected BLL impacts from diet to the estimated average BLLs for LMICs and HICs. The best available estimates for regional and national average BLLs are from IHME ^22^, however, these likely have a substantial level of uncertainty. With the scarcity of nationally representative data for BLLs in LMICs, IHME’s estimates rely on covariates relating to socio-demographics and time since leaded gasoline phase-out to predict national BLLs. Specifically, IHME’s modelling does not currently consider the available TDS data for dietary lead intake. Meanwhile, since the phase-out of leaded-fuels, food crop contamination is suggested to have increased in some regions due to increased industrial lead pollution ^16^. Therefore, disagreements between IHME estimates for national BLLs and the BLL impacts from food should be expected, particularly in countries where food contamination is expected to be high and BLL data is limited. Hence, the uncertainties in the estimated average BLLs in LMICs are brought forward into our estimation of the % of BLLs attributable to dietary lead intake.

## 5. Conclusion

This study estimates the contribution of dietary lead intake to global BLLs, often overlooked as a key driver of the lead poisoning health burden. Dietary lead intake data is taken from 25 total diet studies (TDS) across 46 countries. For children, the population-weighted average dietary lead intake in LMICs (32.0 μg/day) is found to be more than three times higher than in HICs (9.3 μg/day), and more than 10 times higher than the FDA’s reference level for children (2.2 μg/day). Vegetables (27%) and cereals (24%) contributed the most to dietary lead intake across the TDS, and lead uptake into food crops from the environment is suggested to be a primary driver of food contamination and dietary lead intake. The population-weighted mean BLL impacts predicted by the AALM for children are 29 μg/L in LMICs and 12 μg/L in HICs. The AALM is suggested to potentially overestimate BLL impact for adults (61 μg/L in LMICs), although more conservative methods for predicting BLL impacts still show high BLL impact with an upper and lower bound range of 17-53 μg/L from diet for adults in LMICs and 5-14 μg/L in HICs.

Compared to estimates for average BLLs in LMICs and HICs, dietary lead intake is expected to potentially be responsible for 40-50% of BLLs in LMICs, and potentially more than 50% of BLLs in HICs. These results suggest that dietary lead intake is likely a major global driver of BLLs, in line with previous statements from the WHO and EFSA (EFSA, 2010; WHO, 2007). However, dietary lead intake and the associated BLL impacts are expected to vary significantly between countries and regions. The burden of dietary lead intake is expected to be concentrated in South Asia, in line with prior literature that suggests South Asia as the global epicentre for both lead poisoning and food contamination ^16,22^.

The accuracy of the results is limited by uncertainties relating to the TDS data, biokinetic modelling, and IHME estimates for average BLLs. Nonetheless, the available data suggests that the magnitude of the health burden of lead exposure from dietary lead intake is substantial. Furthermore, current regulatory levels for maximum lead concentrations in foods (0.05-0.3 mg/kg) are found to be inadequate, implying a daily dietary lead intake of approximately 200 μg/day, far higher than the FDA reference level (2.2 μg/day). Therefore, research and interventions to reveal and address the health burden of dietary lead intake should be a priority. Efforts are needed to revise regulatory levels for maximum lead concentrations in foods, conduct more TDS studies in LMICs, and investigate the key drivers of dietary lead intake and food contamination.

## Data Availability

All data produced in the present study are available upon reasonable request to the authors

## Acknowledgements

The authors would like to acknowledge the support of Pure Earth’s Innovation Lab for facilitating this research.

## References

1 IHME. GBD Results Tool. 2025.https://vizhub.healthdata.org/gbd-results/ (accessed 8 Dec2025).

2 Larsen B, Sánchez-Triana E. Global health burden and cost of lead exposure in children and adults: a health impact and economic modelling analysis. Lancet Planet Health 2023; 7: e831–e840.

3 Kinally C, Fuller R, Larsen B, Hu H, Lanphear B. A review of lead exposure source attributional studies. Science of the Total Environment. 2025; 990. doi:10.1016/j.scitotenv.2025.179838.

4 ATSDR. Toxicological Profile for Lead. 2020.

5 WHO. Health risks of heavy metals from long-range transboundary air pollution. World Health Organization Regional Office Europe, 2007.

6 EFSA. Scientific Opinion on Lead in Food. EFSA Journal 2010; 8.

7 JECFA. Summary and conclusions of the 53rd meeting of the Joint FAO/WHO Expert Committee on Food Additives. 1999.

8 WHO. Evaluation of certain food additives and contaminants: seventy-third report of the Joint FAO/WHO Expert Committee on Food Additives. WHO technical report series; 960. 2011.

9 Flannery BM, Middleton KB. Updated interim reference levels for dietary lead to support FDA’s Closer to Zero action plan. Regulatory Toxicology and Pharmacology. 2022; 133. doi:10.1016/j.yrtph.2022.105202.

10 Carrington C, Bolger PM. An Assessment of the Hazards of Lead in Food. Regul Toxicol Pharmacol 1992; 16.

11 JECFA. WHO Food Additives Series: 64 Safety evaluation of certain food additives and contaminants. Prepared by the Seventy-third meeting of the Joint FAO/ WHO Expert Committee on Food Additives (JECFA). Lead. 2011.

12 US EPA. Technical Support Document for the All Ages Lead Model (AALM) version 3.0 – Parameters, Equations, and Evaluations. 2024.

13 Gergelová P, Martino L, Rovesti E. EFSA scientific report on dietary exposure to lead in the European population. EFSA Journal 2025; 23. doi:10.2903/j.efsa.2025.9577.

14 Zhao X, Shao Y, Ma L, Shang X, Zhao Y, Wu Y. Exposure to Lead and Cadmium in the Sixth Total Diet Study-China, 2016−2019. China CDC Wkly 2022; 4.

15 Carrington C, Devleesschauwer B, Gibb HJ, Bolger PM. Global burden of intellectual disability resulting from dietary exposure to lead, 2015. Environ Res 2019; 172: 420–429.

16 Xiao N, Wang Q, Wang Y, Yao Y, Shen C. Four decades of global cereal lead contamination patterns. Ecotoxicol. Environ. Saf. 2026; 309. doi:10.1016/j.ecoenv.2026.119687.

17 Vasconcelos Neto MC de, Silva TBC, Araújo VE de, Souza SVC de. Lead contamination in food consumed and produced in Brazil: Systematic review and meta-analysis. Food Research International 2019; 126. doi:10.1016/j.foodres.2019.108671.

18 Huang X, Zhao B, Wu Y, Tan M, Shen L, Feng G et al. The lead and cadmium content in rice and risk to human health in China: A systematic review and meta-analysis. PLoS One. 2022; 17. doi:10.1371/journal.pone.0278686.

19 Codex Alimentarius. Guidelines for the Simple Evaluation of Dietary Exposure to Food AdditivesCAC/GL 3-1989. 2014.

20 Gimou MM, Pouillot R, Charrondiere UR, Noël L, Guérin T, Leblanc JC. Dietary exposure and health risk assessment for 14 toxic and essential trace elements in Yaoundé: The Cameroonian total diet study. Food Additives and Contaminants - Part A 2014; 31: 1064–1080.

21 Fuller R, Porterfield K, Hanrahan D, Hu H. Cumulative population blood lead levels. BMJ Glob. Health. 2025; 10. doi:10.1136/bmjgh-2024-018145.

22 IHME. Global Burden of Disease Collaborative Network. Global Burden of Disease Study 2023 (GBD 2023) Lead Exposure Estimates 1990-2023. Seattle, United States of America, 2025.

23 Spungen JH. Children’s exposures to lead and cadmium: FDA total diet study 2014-16. Food Addit Contam Part A Chem Anal Control Expo Risk Assess 2019; 36: 893–903.

24 Rossi MC, Castanheira I, Sammán NC. Lead, cadmium and arsenic exposure of schoolchildren of northwest Argentina from a risk assessment study. Food Addit Contam Part A Chem Anal Control Expo Risk Assess 2019; 36: 1314–1326.

25 Kinally C, Sefton E, Chang C, Hu H, Fuller R. Understanding the anthropogenic sources and pathways of lead contamination of food crops: A review of current research and research needs. under review 2026.

26 Surawi NH, Nurul Neswatul Hasanah Noor Azhar, Siti Irfah Ishak, Laila Rabaah Ahmad Suhaimi, Norzitah Abu Khair, Zailina Abdul Majid. Malaysian Total Diet Study (TDS) 2019/2020. 2025.

27 Islam MS, Ahmed MK, Habibullah-Al-Mamun M, Islam KN, Ibrahim M, Masunaga S. Arsenic and lead in foods: a potential threat to human health in Bangladesh. Food Addit Contam Part A Chem Anal Control Expo Risk Assess 2014; 31: 1982–1992.

28 Sharma M, Maheshwari M, Morisawa S. Dietary and inhalation intake of lead and estimation of blood lead levels in adults and children in Kanpur, India. Risk Analysis 2005; 25: 1573–1588.

29 González-Pérez de Medina L, Muñoz-Fariña O, Fernández-Guerrero Y, Roman-Benn A, Bastias-Montes JM, Quevedo-León R et al. Arsenic, lead and cadmium concentration in food and estimated daily intake in the Cuban population and the health risks using a Total Diet Study. J Environ Sci Health B 2024; 59: 112–122.

30 Neto MV, Quintal APN, Pôrto LBG, Vitorino Carvalho de Souza S. Lead in Brazilian food: Exposure assessment and risk characterization. Food Addit Contam Part A Chem Anal Control Expo Risk Assess 2021; 38: 315–325.

31 Codex Alimentarius. The Code of Practice for the Prevention and Reduction of Lead Contamination in Foods (CXC 56-2004). 2004.

32 Kumar A, Kumar A, Cabral-Pinto M, Chaturvedi AK, Shabnam AA, Subrahmanyam G et al. Lead toxicity: Health hazards, influence on food Chain, and sustainable remediation approaches. Int. J. Environ. Res. Public Health. 2020; 17. doi:10.3390/ijerph17072179.

33 Malavolti M, Fairweather-Tait SJ, Malagoli C, Vescovi L, Vinceti M, Filippini T. Lead exposure in an Italian population: Food content, dietary intake and risk assessment. Food Research International 2020; 137. doi:10.1016/j.foodres.2020.109370.

34 Lee JW, Choi H, Hwang UK, Kang JC, Kang YJ, Kim K Il et al. Toxic effects of lead exposure on bioaccumulation, oxidative stress, neurotoxicity, and immune responses in fish: A review. Environ. Toxicol. Pharmacol. 2019; 68: 101–108.

35 Vasconcelos Neto M, Quintal APN, Pôrto LBG, Vitorino Carvalho de Souza S. Lead in Brazilian food: Exposure assessment and risk characterization. Food Addit Contam Part A Chem Anal Control Expo Risk Assess 2021; 38: 315–325.

36 Bischoff K, Thompson B, Erb HN, Higgins WP, Ebel JG, Hillebrandt JR. Declines in blood lead concentrations in clinically affected and unaffected cattle accidentally exposed to lead. Journal of Veterinary Diagnostic Investigation 2012; 24: 182–187.

37 Ericson B, Hu H, Nash E, Ferraro G, Sinitsky J, Taylor MP. Blood lead levels in low-income and middle-income countries: a systematic review. Lancet Planet Health 2021; 5: e145–e153.

38 Shamsudin SB, Marzuki A, Jeffree MS, Awang Lukman K. BLOOD LEAD CONCENTRATION AND WORKING MEMORY ABILITY ON MALAY PRIMARY SCHOOL CHILDREN IN URBAN AND RURAL AREA, MALACCA. Acta Scientifica Malaysia 2017; 1: 4–7.

39. FAO/WHO Codex Alimentarius. Codex Alimentarius Commission (2025) General Standard for Contaminants and Toxins in Food and Feed (CXS 193-1995). 2025.

40 EU. Commission Regulation (EU) 2023/915 of 25 April 2023 on maximum levels for certain contaminants in food and repealing Regulation (EC) No 1881/2006. 2023.

41 Health Canada. Health Canada’s Maximum Levels for Chemical Contaminants in Foods. 2020.https://www.canada.ca/en/health-canada/services/food-nutrition/food-safety/chemical-contaminants/maximum-levels-chemical-contaminants-foods.html?utm_source=chatgpt.com (accessed 18 Dec2025).

42 NHC & SAMR. National Health Commission of the People’s Republic of China & State Administration for Market Regulation (2022) National Food Safety Standard—Maximum Levels of Contaminants in Foods (GB 2762-2022). 2022.

43 FSSAI. Food Safety and Standards Authority of India (FSSAI) (2011) Food Safety and Standards (Contaminants, Toxins and Residues) Regulations, 2011 (as updated/compiled PDF). 2011.

44 Utembe W, Gulumian M. Issues and challenges in the application of the ieubk model in the health risk assessment of lead: A case study from blantyre Malawi. Int J Environ Res Public Health 2021; 18. doi:10.3390/ijerph18158207.

45 Kordas K. The ‘Lead Diet’: Can Dietary Approaches Prevent or Treat Lead Exposure? J Pediatr 2017; 185: 224–231.e1.

