## Supporting information for "Quantifying the global burden of lead exposure from dietary lead intake"

East Asia

South Asia

LAC

Sub-Saharan Africa

Middle East & North Africa

Europe, North America and Pacific

|  | Lead intake g /day |
| --- | --- |
| Malaysia (Surawi et al., 2025) | 51.302 |
| Japan (Watanabe et al. 2022) | 3.842 |
| China (Zhao et al., 2022) | 6.554 |
| South Korea (Lee et al. 2006) | 28.928 |
| Bangladesh (Islam et al., 2014) | 158.2 |
| Kanpur India (Sharma et al., 2005) | 41.9456 |
| Cuba (González-Pérez de Medina et al., 2024) | 44.635 |
| Brazil (Vasconcelos Neto et al., 2021) | 34.578 |
| Argentina (Rossi et al., 2019) | 13.56 |
| Nigeria (Ingenbleek et al., 2020) | 21.696 |
| Mali (Ingenbleek et al., 2020) | 6.554 |
| Cameroon (Ingenbleek et al., 2020) | 7.006 |
| Benin (Ingenbleek et al., 2020) | 9.04 |
| Yaoundé, Cameroon (Gimou et al. 2014) | 19.8202 |
| Iran (Heshmati et al., 2020) | 25.2216 |
| Riyadh, Saudi Arabia (Othman, 2010) | 7.9326 |
| Beirut, Lebanon (Nasreddine et al., 2006) | 4.8138 |
| EU (Gergelová et al., 2025) | 17.176 |
| Italy (Malavolti et al., 2020) | 3.503 |
| Ireland (FSAI, 2016) | 2.373 |
| Germany (Schneider et al., 2014) | 11.6164 |
| France (Arnich et al. 2012) | 5.424 |
| UK (Rose et al. 2010) | 3.164 |
| US (Spungen, 2019) | 2.599 |
| New Zealand (Pearson et al., 2018) | 2.00171 |

**Figure S1.** Dietary lead intake (μg /day) for a 7-year-old 22.6 kg child. Solid fill = national data, pattern fill = sub-national datasets, green = child specific dietary lead intake data. LAC = Latin America & Caribbean

g/day

|  | Malaysia (Surawi et al., 2025) | EU (EFSA, 2025) | Cuba (González-Pérez de Medina et al., 2024) | China (Zhao et al., 2022) | Brazil (Vasconcelos Neto et al., 2021) | Italy (Malavolti et al., 2020) | Brazil (Vasconcelos Neto et al., 2019) | Ireland (FSAI, 2016) | Yaoundé, Cameroon (Gimou et al., 2014) | Germany (K. Schneider et al., 2014) | China (Yingliang et al., 2014) | Bangladesh (Islam et al., 2014) | France (Arnich et al., 2012) | UK (Rose et al., 2010) | Riyadh, Saudi Arabia (Othman, 2010) | Lebanon (Nasreddine et al., 2006) | Kanpur, India (Sharma et al., 2005) |
| --- | --- | --- | --- | --- | --- | --- | --- | --- | --- | --- | --- | --- | --- | --- | --- | --- | --- |
| Cereals | 68.31 | 2.37687 | 17.1151 | 7.569 | 3.9913 | 1.767 | 14.4097 | 0.528 | 3.1572 | 4.6813 | 34.0032 | 24.5857 | 1.944 | 1.68 | 5.09652 | 6.9012 | 60 |
| Sugar products | 0 | 2.34318 | 2.48449 | 0 | 0 | 0.558 | 0.472449 | 0 | 0 | 0 | 0 | 0 | 0.432 | 0.3 | 1.72692 | 0 | 0 |
| Fats & oils | 0 | 0.200942 | 15.0549 | 0 | 0 | 0 | 0.472449 | 0 | 0 | 0.34194 | 0 | 0 | 0 | 0.12 | 0.12636 | 0 | 0 |
| Vegetables | 10.35 | 2.00607 | 37.4849 | 5.046 | 53.7291 | 2.79 | 36.3786 | 0.288 | 21.048 | 5.22527 | 27.3504 | 124.05 | 1.404 | 1.56 | 6.90768 | 2.6838 | 30.225 |
| Fruits | 0 | 0.910656 | 7.25667 | 0 | 0.92107 | 0.558 | 0 | 0.192 | 0 | 4.68731 | 0.7392 | 7.376 | 0.324 | 0.42 | 1.13724 | 1.5336 | 0 |
| Beverages |  | 1.5631 | 9.11232 | 1.566 | 21.7987 | 1.767 | 19.6066 | 0.84 | 4.7358 | 7.69838 | 0 | 0 | 1.188 | 1.02 | 2.04282 | 0 | 0 |
| Meat | 4.14 | 3.6573 | 7.03855 | 0 | 1.84214 | 0.93 | 16.5357 | 0.048 | 0 | 2.54164 | 6.6528 | 0 | 0.432 | 0.42 | 2.71674 | 0.639 | 0 |
| Fish & seafood | 0 | 0.182846 | 2.31578 | 0 | 0 | 0.372 | 0 | 0.048 | 3.6834 | 0.369768 | 2.9568 | 18.7549 | 0.216 | 0.06 | 0 | 0 | 0 |
| Eggs | 0 | 0.0205198 | 0.610048 | 0 | 0 | 0 | 0.472449 | 0 | 0 | 0.13944 | 1.4784 | 1.844 | 0 | 0.06 | 0 | 0 | 0 |
| Milk & dairy | 0 | 0.69081 | 12.5452 | 0 | 0 | 0.372 | 6.14184 | 0.096 | 0 | 4.35482 | 0.7392 | 7.2118 | 2.16 | 0.36 | 1.07406 | 0.97128 | 2.925 |

**Figure S2**. Total dietary lead intake by food group (μg/day) for the 18 TDS studies which provide data by food group. Additional meta-analysis studies included for China 2014 and Brazil 2019, not included in the BLL impacts assessment (superseded by government commissioned national TDS). All beverages = alcoholic and non-alcoholic beverages, excluding tap water.

Blood lead level (g /L)

| Age (years) | AALM male | IEUBK |
| --- | --- | --- |
| 1 | 17.69 | 19 |
| 1.003 | 17.73 | 19 |
| 1.005 | 17.77 | 19 |
| 1.008 | 17.82 | 19 |
| 1.011 | 17.86 | 19 |
| 1.014 | 17.9 | 19 |
| 1.016 | 17.94 | 19 |
| 1.019 | 17.975 | 19 |
| 1.022 | 18.015 | 19 |
| 1.025 | 18.045 | 19 |
| 1.027 | 18.08 | 19 |
| 1.03 | 18.115 | 19 |
| 1.033 | 18.145 | 19 |
| 1.036 | 18.175 | 19 |
| 1.038 | 18.21 | 19 |
| 1.041 | 18.235 | 19 |
| 1.044 | 18.26 | 19 |
| 1.047 | 18.29 | 19 |
| 1.049 | 18.32 | 19 |
| 1.052 | 18.345 | 19 |
| 1.055 | 18.37 | 19 |
| 1.058 | 18.395 | 19 |
| 1.06 | 18.425 | 19 |
| 1.063 | 18.445 | 19 |
| 1.066 | 18.465 | 19 |
| 1.068 | 18.495 | 19 |
| 1.071 | 18.515 | 19 |
| 1.074 | 18.54 | 19 |
| 1.077 | 18.56 | 19 |
| 1.079 | 18.58 | 19 |
| 1.082 | 18.605 | 19 |
| 1.085 | 18.625 | 19 |
| 1.088 | 18.65 | 19 |
| 1.09 | 18.67 | 19 |
| 1.093 | 18.69 | 19 |
| 1.096 | 18.71 | 19 |
| 1.099 | 18.73 | 19 |
| 1.101 | 18.75 | 19 |
| 1.104 | 18.77 | 19 |
| 1.107 | 18.785 | 19 |
| 1.11 | 18.805 | 19 |
| 1.112 | 18.825 | 19 |
| 1.115 | 18.845 | 19 |
| 1.118 | 18.865 | 19 |
| 1.121 | 18.885 | 19 |
| 1.123 | 18.895 | 19 |
| 1.126 | 18.915 | 19 |
| 1.129 | 18.935 | 19 |
| 1.132 | 18.955 | 19 |
| 1.134 | 18.97 | 19 |
| 1.137 | 18.99 | 19 |
| 1.14 | 19.005 | 19 |
| 1.142 | 19.025 | 19 |
| 1.145 | 19.04 | 19 |
| 1.148 | 19.06 | 19 |
| 1.151 | 19.08 | 19 |
| 1.153 | 19.09 | 19 |
| 1.156 | 19.11 | 19 |
| 1.159 | 19.13 | 19 |
| 1.162 | 19.14 | 19 |
| 1.164 | 19.16 | 19 |
| 1.167 | 19.18 | 19 |
| 1.17 | 19.195 | 19 |
| 1.173 | 19.21 | 19 |
| 1.175 | 19.225 | 19 |
| 1.178 | 19.245 | 19 |
| 1.181 | 19.26 | 19 |
| 1.184 | 19.275 | 19 |
| 1.186 | 19.295 | 19 |
| 1.189 | 19.305 | 19 |
| 1.192 | 19.325 | 19 |
| 1.195 | 19.335 | 19 |
| 1.197 | 19.355 | 19 |
| 1.2 | 19.375 | 19 |
| 1.203 | 19.385 | 19 |
| 1.205 | 19.405 | 19 |
| 1.208 | 19.415 | 19 |
| 1.211 | 19.435 | 19 |
| 1.214 | 19.45 | 19 |
| 1.216 | 19.465 | 19 |
| 1.219 | 19.48 | 19 |
| 1.222 | 19.495 | 19 |
| 1.225 | 19.51 | 19 |
| 1.227 | 19.53 | 19 |
| 1.23 | 19.54 | 19 |
| 1.233 | 19.56 | 19 |
| 1.236 | 19.57 | 19 |
| 1.238 | 19.59 | 19 |
| 1.241 | 19.6 | 19 |
| 1.244 | 19.62 | 19 |
| 1.247 | 19.63 | 19 |
| 1.249 | 19.65 | 19 |
| 1.252 | 19.66 | 19 |
| 1.255 | 19.68 | 19 |
| 1.258 | 19.695 | 19 |
| 1.26 | 19.71 | 19 |
| 1.263 | 19.725 | 19 |
| 1.266 | 19.735 | 19 |
| 1.268 | 19.755 | 19 |
| 1.271 | 19.765 | 19 |
| 1.274 | 19.785 | 19 |
| 1.277 | 19.795 | 19 |
| 1.279 | 19.815 | 19 |
| 1.282 | 19.825 | 19 |
| 1.285 | 19.84 | 19 |
| 1.288 | 19.855 | 19 |
| 1.29 | 19.87 | 19 |
| 1.293 | 19.885 | 19 |
| 1.296 | 19.895 | 19 |
| 1.299 | 19.915 | 19 |
| 1.301 | 19.925 | 19 |
| 1.304 | 19.94 | 19 |
| 1.307 | 19.955 | 19 |
| 1.31 | 19.97 | 19 |
| 1.312 | 19.985 | 19 |
| 1.315 | 20 | 19 |
| 1.318 | 20.01 | 19 |
| 1.321 | 20.03 | 19 |
| 1.323 | 20.04 | 19 |
| 1.326 | 20.055 | 19 |
| 1.329 | 20.07 | 19 |
| 1.332 | 20.08 | 19 |
| 1.334 | 20.1 | 19 |
| 1.337 | 20.11 | 19 |
| 1.34 | 20.125 | 19 |
| 1.342 | 20.14 | 19 |
| 1.345 | 20.15 | 19 |
| 1.348 | 20.165 | 19 |
| 1.351 | 20.18 | 19 |
| 1.353 | 20.195 | 19 |
| 1.356 | 20.21 | 19 |
| 1.359 | 20.225 | 19 |
| 1.362 | 20.235 | 19 |
| 1.364 | 20.25 | 19 |
| 1.367 | 20.265 | 19 |
| 1.37 | 20.275 | 19 |
| 1.373 | 20.295 | 19 |
| 1.375 | 20.305 | 19 |
| 1.378 | 20.315 | 19 |
| 1.381 | 20.335 | 19 |
| 1.384 | 20.345 | 19 |
| 1.386 | 20.355 | 19 |
| 1.389 | 20.375 | 19 |
| 1.392 | 20.385 | 19 |
| 1.395 | 20.4 | 19 |
| 1.397 | 20.41 | 19 |
| 1.4 | 20.425 | 19 |
| 1.403 | 20.44 | 19 |
| 1.405 | 20.45 | 19 |
| 1.408 | 20.465 | 19 |
| 1.411 | 20.48 | 19 |
| 1.414 | 20.49 | 19 |
| 1.416 | 20.505 | 19 |
| 1.419 | 20.52 | 19 |
| 1.422 | 20.53 | 19 |
| 1.425 | 20.55 | 19 |
| 1.427 | 20.56 | 19 |
| 1.43 | 20.57 | 19 |
| 1.433 | 20.585 | 19 |
| 1.436 | 20.6 | 19 |
| 1.438 | 20.61 | 19 |
| 1.441 | 20.625 | 19 |
| 1.444 | 20.635 | 19 |
| 1.447 | 20.65 | 19 |
| 1.449 | 20.665 | 19 |
| 1.452 | 20.675 | 19 |
| 1.455 | 20.69 | 19 |
| 1.458 | 20.7 | 19 |
| 1.46 | 20.715 | 19 |
| 1.463 | 20.725 | 19 |
| 1.466 | 20.74 | 19 |
| 1.468 | 20.755 | 19 |
| 1.471 | 20.765 | 19 |
| 1.474 | 20.775 | 19 |
| 1.477 | 20.795 | 19 |
| 1.479 | 20.805 | 19 |
| 1.482 | 20.815 | 19 |
| 1.485 | 20.83 | 19 |
| 1.488 | 20.845 | 19 |
| 1.49 | 20.855 | 19 |
| 1.493 | 20.865 | 19 |
| 1.496 | 20.88 | 19 |
| 1.499 | 20.895 | 19 |
| 1.501 | 20.905 | 19 |
| 1.504 | 20.92 | 19 |
| 1.507 | 20.93 | 19 |
| 1.51 | 20.94 | 19 |
| 1.512 | 20.955 | 19 |
| 1.515 | 20.97 | 19 |
| 1.518 | 20.98 | 19 |
| 1.521 | 20.99 | 19 |
| 1.523 | 21 | 19 |
| 1.526 | 21.02 | 19 |
| 1.529 | 21.03 | 19 |
| 1.532 | 21.04 | 19 |
| 1.534 | 21.05 | 19 |
| 1.537 | 21.07 | 19 |
| 1.54 | 21.08 | 19 |
| 1.542 | 21.09 | 19 |
| 1.545 | 21.1 | 19 |

| Age (years) | JECFA lower bound | JECFA upper bound |
| --- | --- | --- |
| 1 | 5.8745 | 18.7984 |
| 2 | 7.45367 | 23.8517 |
| 3 | 8.9065 | 28.5008 |
| 4 | 10.233 | 32.7456 |
| 5 | 11.5595 | 36.9904 |
| 6 | 12.8228 | 41.0331 |

| Age (years) | JECFA adult lower bound | JECFA adult upper bound | FDA | JECFA midpoint |
| --- | --- | --- | --- | --- |
| 16 | 14.1215 | 42.9786 | 24.5592 | 28.5501 |
| 17 | 14.9642 | 45.5432 | 26.0247 | 30.2537 |
| 18 | 15.7778 | 48.0193 | 27.4396 | 31.8985 |
| 19 | 16.6204 | 50.5839 | 28.9051 | 33.6021 |
| 20 | 17.434 | 53.06 | 30.32 | 35.247 |
| 21 | 17.434 | 53.06 | 30.32 | 35.247 |
| 22 | 17.434 | 53.06 | 30.32 | 35.247 |
| 23 | 17.434 | 53.06 | 30.32 | 35.247 |
| 24 | 17.434 | 53.06 | 30.32 | 35.247 |
| 25 | 17.434 | 53.06 | 30.32 | 35.247 |

| Age (years) | JECFA midpoint |
| --- | --- |
| 1 | 12.3364 |
| 2 | 15.6527 |
| 3 | 18.7036 |
| 4 | 21.4893 |
| 5 | 24.275 |
| 6 | 26.9279 |
|  | 29.9789 |

**Figure S3**. Comparison of estimates for the dose response relationship between dietary lead intake and blood lead level from ages 1-25. BLL estimates consider a dietary lead intake of 1.05 μg/kgBW/day, using the All Ages Lead Model (AALM), the IEUBK model, and the linear dietary lead intake to BLL conversion factors from FDA and JECFA for children (0-6 years old) and adults (>16 years old). AALM BLL impact is an average of BLLs predicted for males and females, IEUBK, FDA and JECFA do not differentiate by sex.

East Asia

South Asia

LAC

Sub-Saharan Africa

Middle East & North Africa

Europe, North America and Pacific

Blood lead level impact g /L

|  | Food AALM | IHME national average |
| --- | --- | --- |
| Malaysia (Surawi et al., 2025) | 49.13 | 7.7 |
| Japan (Watanabe et al. 2022) | 3.685 | 15.2 |
| China (Zhao et al., 2022) | 6.285 | 30.6 |
| South Korea (Lee et al. 2006) | 27.71 | 26 |
| Bangladesh (Islam et al., 2014) | 141.1 | 85.6 |
| Kanpur India (Sharma et al., 2005) | 40.175 | 77.7 |
| Cuba (González-Pérez de Medina et al., 2024) | 42.755 | 54 |
| Brazil (Vasconcelos Neto et al., 2021) | 33.12 | 24.8 |
| Argentina (Rossi et al., 2019) | 12.995 | 28.2 |
| Nigeria (Ingenbleek et al., 2020) | 11.785 | 45.1 |
| Mali (Ingenbleek et al., 2020) | 6.285 | 83 |
| Cameroon (Ingenbleek et al., 2020) | 6.715 | 99.9 |
| Benin (Ingenbleek et al., 2020) | 8.665 | 73.2 |
| Yaoundé, Cameroon (Gimou et al. 2014) | 12.325 | 102.9 |
| Iran (Heshmati et al., 2020) | 24.155 | 35.8 |
| Riyadh, Saudi Arabia (Othman, 2010) | 7.605 | 33.6 |
| Beirut, Lebanon (Nasreddine et al., 2006) | 4.615 | 38.3 |
| EU (Gergelová et al., 2025) | 23.885 | 22.6 |
| Italy (Malavolti et al., 2020) | 3.36 | 11.9 |
| Ireland (FSAI, 2016) | 2.28 | 16.6 |
| Germany (Schneider et al., 2014) | 11.13 | 13.4 |
| France (Arnich et al. 2012) | 5.2 | 15.8 |
| UK (Rose et al. 2010) | 3.43 | 14 |
| US (Spungen, 2019) | 3.01 | 10.8 |
| New Zealand (Pearson et al., 2018) | 5.35 | 21.3 |

**Figure S4.** Blood lead level impacts (μg /dl) for a 7-year-old 22.6 kg child. Pattern fill = sub-national datasets, green = child specific dietary lead intake data, grey = IHME estimated national average BLL for year of TDS publication, for TDS published after 2023 IHME 2023 (latest available) data is used. LAC = Latin America & Caribbean.
